# A simulation-based procedure to estimate base rates from Covid-19 antibody test results I: Deterministic test reliabilities

**DOI:** 10.1101/2020.04.28.20075036

**Authors:** Reinoud Joosten, Abhishta Abhishta

## Abstract

We design a procedure (*the complete Python code may be obtained at* https://github.com/abhishta91/antibody_montecarlo) using Monte Carlo (MC) simulation to establish the point estimators described below and confidence intervals for the base rate of occurence of an attribute (e.g., antibodies against Covid-19) in an aggregate population (e.g., medical care workers) based on a test. The requirements for the procedure are the test’s sample size (*N*) and total number of positives (*X*), and the data on test’s reliability.

The **modus** is the prior which generates the largest frequency of observations in the MC simulation with precisely the number of test positives (maximum-likelihood estimator). The **median** is the upper bound of the set of priors accounting for half of the total relevant observations in the MC simulation with numbers of positives identical to the test’s number of positives.

Our rather preliminary findings are
- The median and the confidence intervals suffice universally.
- The estimator 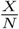 may be outside of the two-sided 95% confidence interval.
- Conditions such that the modus, the median and another promising estimator which takes the reliability of the test into account, are quite close.
- Conditions such that the modus and the latter estimator must be regarded as logically inconsistent.
- Conditions inducing rankings among various estimators relevant for issues concerning over-or underestimation.

**JEL-codes:** C11, C13, C63

## Introduction

The Corona crisis revealed several bottle necks regarding testing. Many of these bottle necks are physical, but one is cognitive: how to interpret the results of a test. Medical experts seem to have problems in interpreting and combining statistical information (cf., e.g., Uffrage *et al*. [2000]). They, as well as politicians, journalists, or the general public, may suffer from the so-called base-rate fallacy (cf., Bar-Hillel [1980]):

> The base-rate fallacy is people’s tendency to ignore base rates in favor of, e.g., individuating information (when such is available), rather than integrate the two. This tendency has important implications for understanding judgment phenomena in many clinical, legal, and social-psychological settings.

The *base rate* in the quote above can be associated with the incidence of an attribute in a larger population, such as the occurrence of antibodies against the Corona virus in a certain region or profession, breast cancer among females, or Down syndrome among unborn children with mothers aged 41. The *individuating information* in the quote above can be associated with information obtained from a(n individual) test (result).

A widely accepted technique integrating the two kinds of information mentioned, involves Bayesian reasoning in which a prior distribution (base rate) is updated on the basis of information gained from a (possibly imperfect) test, such that the latter can be interpreted on an individual level. It safe to say that this technique is not very well known throughout the various scientific communities, let alone to the general public. It is also safe to say that the technique yields counter-intuitive answers. There are at least two sides to this science-versus-intuition gap: on the one hand human intuition seems underrated and should be taken more seriously, and on the other intuition can be helped by representing statistical data in a more user friendly manner (cf., e.g., Cosmides & Tooby [1996], Gigerenzer & Hoffrage [1995]).

The following stylized problem has been used recently for didactic purposes to inform the general public about the limited use of testing in case the general population has a low incidence of an attribute (cf., Volkskrant [2020]).

**Example 1**. A test for antibodies against Corona (Covid-19) has the following reliability: if a person really has antibodies, the test gives a positive result with 75%, hence the test gives a negative result with the complementary probability, i.e., 25%; if a person really does not have antibodies, the test gives a negative result with 95% probability, hence the test gives a positive result with the complementary probability of 5%. This information can be summarized as follows:

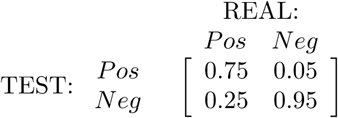

The number 0.05 is also known as the rate of false positives (a.k.a. type I error rate), and the number 0.25 is known as the rate of false negatives (a.k.a. type II error rate).

Now, suppose 2% of the general public have antibodies against Corona. This is the **base rate** (a.k.a. prior in statistical jargon), and we test 10,000 people taking all these probabilities mentioned as given and (exactly) true. Then, the following natural question arises.

- How many people will test positive (in expectation)?

If 10,000 people are tested, then approximately^1^ 200 will have antibodies for real, and the complementary number 9800 will not. Of the approximately 200 people that really have antibodies, approximately 150 test positive, but approximately 50 test negative and this evaluation is incorrect. Of the approximately 9800 people that in reality do not have antibodies, approximately 490 test positive incorrectly, whereas approximately 9310 test negative. We again put up a matrix helping us to visualize this information.

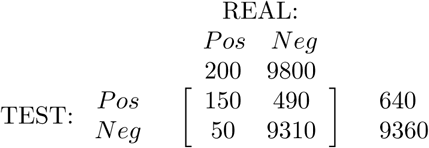

The two numbers above the matrix represent the expected number of people who have antibodies against Corona (left) and those who do not (right). These numbers may be recovered from the matrix below by adding the numbers in the corresponding columns. The two numbers to the right of the matrix represent the expected numbers of people who receive a positive test result, i.e., 640, and a negative one, i.e., 9360. These numbers are obtained by adding the numbers in the corresponding row of the matrix.

We now continue with an analysis based on Bayesian reasoning in order to make sense of these numbers, to answer the ensuing natural questions.

- What is the probability that a person truly has antibodies if tested positive?
- What is the probability that a person truly has antibodies if tested negative?

The probability that a person really has antibodies **if tested positive** is approximated by:

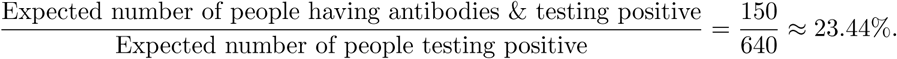

The probability that a person really has antibodies **if tested negative** is approximated by:

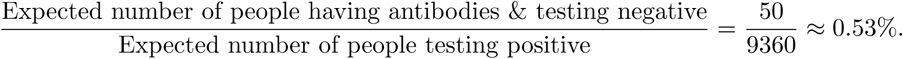

So, receiving a negative test result is rather conclusive as more than 99% of the diagnoses are correct. Receiving a positive test result however, still leaves a lot of room for doubt and insecurity, as the probability that the test result is correct is less than 24%. This means that the vast majority of people receiving a positive test result, receive a misleading diagnosis.

The example above shows that the information gain from a test may be quite disappointing in quality if the incidence levels on a total population level are low. This perceived low quality of information of a positive test result may be a great impediment to promote or justify testing, and it may de-legitimize taking appropriate measures (e.g., wearing face masks, washing hands, forbidding mass meetings or travel), especially if other, non-cognitive, bottle necks occur. For instance, it may be quite costly (reportedly some 45 Euro per test in Robbio^2^ in Italy) or rather time-consuming to test an individual, hence a re-test after a positive test result would be unattractive looking at it from the resource-provision side of the problem, although re-testing in this case will be much, much more informative. An additional bottle neck might be that tests may not be available in sufficient numbers.^3^ Then, a priority or a legitimization problem arises: to use the scarce test for testing people for the first time, or for retesting positives. Especially combinations of these bottle necks, and they have materialized at crucial moments in the Corona crisis, may lead to questioning the usefulness of testing at all.

The aim of this paper is however not to contribute to solving the issue of the base-rate fallacy, nor distributional dilemmas induced by the scarcity of tests. We are interested in solving another bottle neck namely the practical, more basic problem of lack of knowledge (hence unavailability) of a prior distribution (or base rates or incidence rates of occurrence) of an attribute in a chosen aggregate population. We however think there is a psychological connection between the missing base-rate problem and the base-rate fallacy. We suggest that it is very likely that a missing base rate shifts the interpretation of the test’s result unpredictably anywhere between giving a lot of weight (if not all) to the individuating information, or vice versa in which having no anchor for the base rate at all might psychologically mean base rate equals zero.

The reasons why base rates might be lacking can be numerous. Take a Corona test, and suppose that the reliability data were obtained (correctly) in China or Italy, where the illness occurred early and in rather large numbers. If one were to use this test in, for instance, Noord Brabant, the earliest hot spot of Corona in the Netherlands, the validity of the reliability data might be upheld, but the great missing parameter would be the prior, i.e., the incidence of antibodies to the Corona virus on a population level. Assuming the priors to be the same as in Italy or China would be without any scientific base.

An additional aim of this paper is to be able to provide answers regarding priors on the basis of relatively low numbers of tests. Obviously, larger tests provide better answers *if the base rate is stationary*. We have the following reasons for this additional ambition. In case of a disease spreading, the assumption of stationarity is frivolous, so then more is *not* necessarily better, *more recent might be better*. Moreover, crucial measures may be triggered by data on an aggregate level, but cannot be delayed until results from large numbers of tests have accrued. Furthermore, a sequence of estimated priors (using low numbers of tests) taken at different moments in time, may provide information regarding the stationarity issue, in other words: *is it spreading or not?* Additionally, one might have the wish to restrict attention to specific groups each possibly having another base rate, e.g., people working in medical care or care for the elderly, primary school children and teachers, or family members of those working in jobs with a high probability of exposure to Corona.

**Example 2**. We could for instance, use some of the data above to come up with estimates of the probability that antibodies occur in a population. One option is to look at the number of positives which is 640 out of 10,000, but this naive estimate of 6.4% yields a much too high number compared to the real 2% underlying the computations. A seemingly better option is to solve the equation

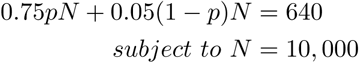

This yields exactly *p* = 0.02 which is the precise prior used for the illustration. So, then we have an estimator, but we have absolutely no idea about how reliable this number is. Let

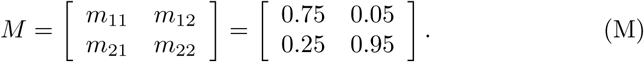

Then it is easy to confirm that the estimator 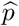 for *p* given the parameters presented, is computed in general terms by

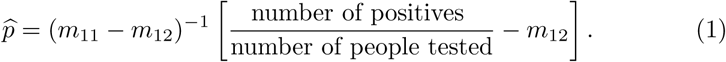

However, even for the given numbers *p* = 0.02*, m*_11_ = 0.75*, m*_12_ = 0.05, to reach this (640) or any given number of positives we have outcomes resulting from a combination of three random processes. Suppose that the number of positives turns out to be 654 instead, which, by the way, may occur with a likelihood quite close to the likelihood of 640 positives occurring, then although the real *p* does not change, its estimator would be 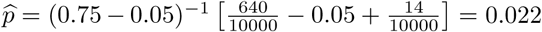.

Observe furthermore that any test result with 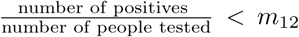 is hard to interpret, or 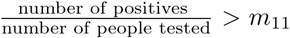 for that matter, because logic dictates that the probability computed should belong to the unit interval.

The organization of the remainder of this note is minimalistic. In the next section, we present results of our Monte Carlo simulations which are used to derive confidence intervals and point estimators for base rates assuming the reliability data to be perfect. The conclusions concentrate on perceived regularities in doing a series of such estimations, and reflections on the feasibility of the aims we started with. The Python codes for anybody wishing to experiment with the tools are available at the github repository.^4^

## 2 Monte Carlo simulation & estimators of priors

We are interested in finding a point estimator or a confidence interval for the base-rate probability of a certain attribute based on a test on this attribute. We operate under the specific assumption that the reliability reported are true. For this purpose we employ the procedure presented in the next subsection. The results for three hypothetical cases are presented and compared. Note that the Monte Carlo simulation can be adapted for many if not all inputs desirable.

### 2.1 Pseudo code

For a certain test (or sample) size *N* meaning the total number of people tested, we find a certain number of positives *X*. A quick approximation using (1):

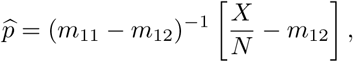

may be convenient to establish a region in the unit interval which base rate qualifies as most likely to underly the statistical process providing the test outcome. In what follows, we make a grid of size 0.001 of the most promising region or interval to be examined more closely. For a given grid size point 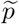 in the latter interval we perform the following loop in pseudo-code.

Step 0 Set *tp*(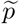*, X, N*) := 0*, K*_1_ := 1*, K*_2_ := 1, 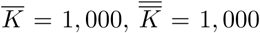. **Go to Step 1**.
Step 1 Draw *N* times with probability 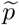 of success to determine^5^ 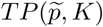. **Go to Step 2**.
Step 2 Draw 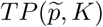 times with probability *m*_11_ of success to determine^6^ 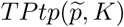. **Go to Step 3**.
Step 3 Draw 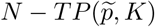 times with probability *m*_22_ of success to determine^7^ 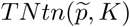, then set^8^ 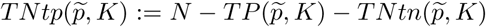. **Go to Step 4**.
Step 4 Set 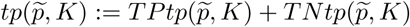. If 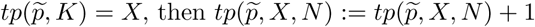. If 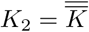, then set *K*_2_ := 1 and set *K*_1_ := *K*_1_ +1 **go to Step 1**. Otherwise if 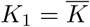, then go to **Step 5**. Otherwise, set *K*_2_ := *K*_2_ + 1 and **go to Step 2**.
Step 5 Save 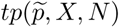.

This sub-loop will run 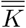 times and larger loop will run 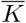 times and register 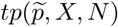 for each such grid point 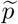, which is simply the number of times in the total Monte Carlo simulation under base rate 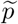, exactly outcome *X* occurs.

### 2.2 Interpretation of results from the MC simulation

By Monte Carlo simulation we generate a large number of positives from a test of size *N* for a fixed known candidate prior which is taken as underlying the simulation, and record how many of the positives out of the total number of positives generated by our Monte Carlo simulation, equal precisely *X*. We rank the, say *G* = 400, candidate priors according to a (n evenly meshed) grid of a relevant interval *p^1^ < p*_2_ *< … < p^G^*.

For candidate prior say *p^j^*, we take, say, 1000 samples of size *N*. For each such sample, we generate a pair consisting of the number of real positives and the number of real negatives by drawing independently *N* observations with probability *p^j^* (1 − *p^j^*) of having (not having) the attribute. Then, for each such pair of numbers, say (*TP, TN*), of true positives and true negatives in the sample, i.e., *TP* + *TN* = *N*, we draw 1000 samples taking *TP* draws with the probability of testing positive equal to the upper left element of *M* and taking *TN* draws with the probability of testing positive equal to the upper right element of *M*. The former are then the *True Positives tested positive* (*TPtp*) and the latter are the *True Negatives tested positive* (*TNtp*).

The sum of those two numbers *TPtp* + *TNtp* then provides one observation of positives 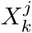. Taking independent samples, we find one million different realizations of positives, say 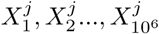. Then, we record among them, the number of positives for known prior *p^j^* being exactly equal to the number of positives resulting from the test as follows

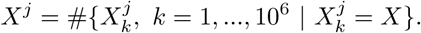

We do the same for the whole range of candidate priors in exactly the same manner.

We then construct a histogram of the *relative* numbers of hits equal to *X* for each prior, i.e.,

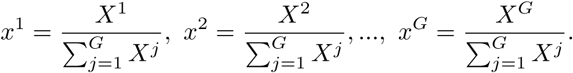

Observe that *x^i^* ≥ 0 for all *i* = 1, 2*, …, G* and that 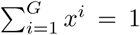. Then, the number *x^i^* tells us that the prior *p^i^* accounted for generating a proportion *x^i^* of all realizations in the entire Monte Carlo simulation yielding *X* positives. So, alternatively these numbers can be interpreted as probabilities. Let in the same vein

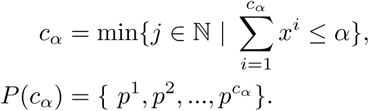

Then, an interpretation for the latter expression immediately comes to mind which is close to the one of a cumulative probability distribution, namely the first *c_α_* of the (ranked) priors that account for proportion of *α* of all realizations in the entire Monte Carlo simulation which yielded exactly *X* positives. The ‘area under the curve’ formed by the histogram between the lower bound of the range examined and *p^cα^*, the latter included, is (approximately) *α*. Continuing along this interesting analogous interpretation we coin the following expressions.

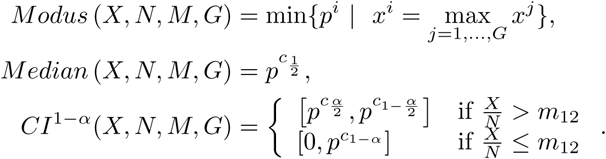

These notions can be interpreted in line with the more standard notions with the same names widely used in statistics.

*Modus* (*X, N, M, G*) is the smallest prior which yields the highest number (proportion) of positives equal to *X* in our Monte Carlo simulation for sample size *N* using *deterministic* reliability matrix *M*, having a grid dividing a relevant interval of priors into *G* parts of equal length. There might be more than one such prior, and in order to obtain a unique prior as *Modus* we took the lowest. So, knowing only little, this prior could be interpreted as a *maximum likelihood estimator* and for the (admittedly few) cases examined we seem to have (with 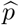 pgiven by Eq. (1)) *Modus* 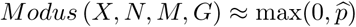. Next, *Median* (*X, N, M, G*) is the smallest prior such that set of priors smaller than or equal to it are responsible for (approximately) half of the simulated hits equal to *X*.

We interpret *CI*^1−α^(*X, N, M, G*) as our confidence interval among the priors as that it gives us the set of priors accounting for a proportion of 1 − *α* of outcomes yielding *X* hits in the Monte Carlo simulation. The restriction in first part of the notion applies to the case that the 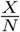 exceeds the type I error rate which intuitively seems a rather convenient turn of events. If the second part applies, i.e., we have a more extreme case of the relative number of hits 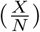 being lower than the type I error rate (*m*_12_), we may obtain with great likelihood *Modus* 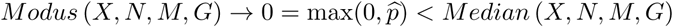.

### 2.3 Results for 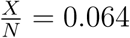*, G* = 400 and *N* ∈ {10^4^, 10^3^, 125}

**Figure 1:**
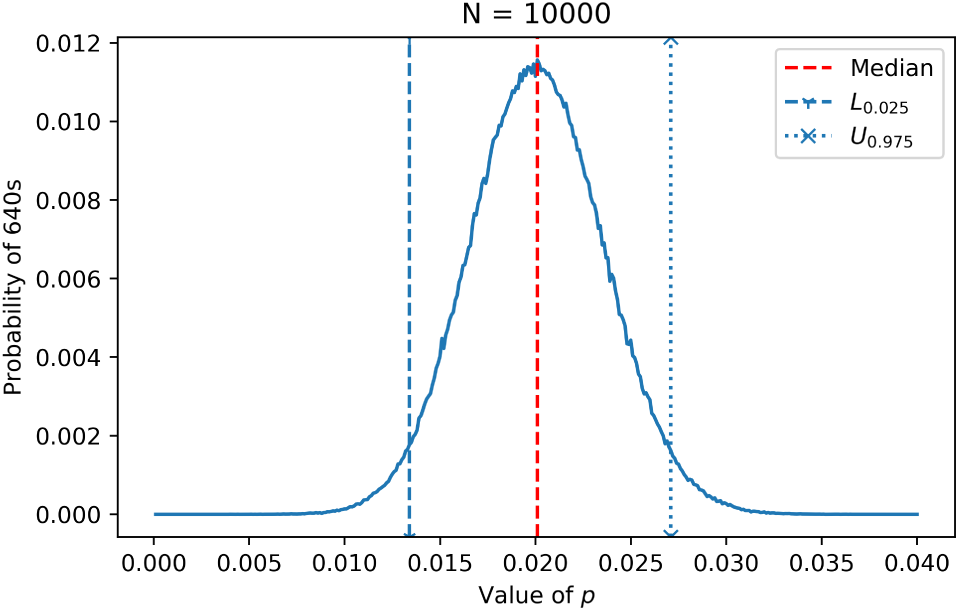
A histogram of the relative number of simulated hits at *X* = 640*, N* = 10, 000 for all values of *p* in the interval [0, 0.04] showing also the location of the median and, the interval of values of *p* in [*L*_0.025_*,U*_0.975_] = [0.0134, 0.0271] responsible for 95% of the simulated hits for *X*.

**Figure 2:**
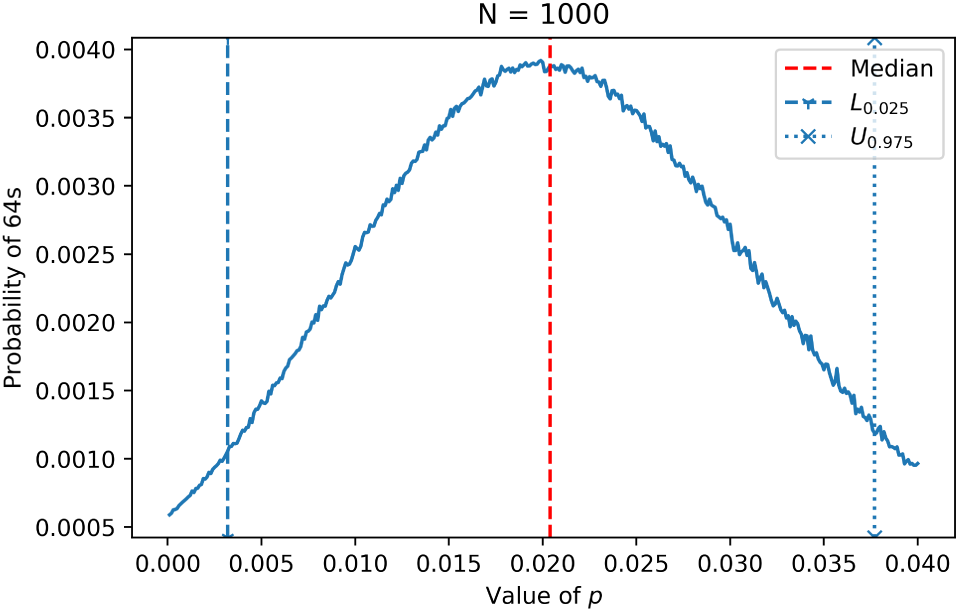
A histogram of the relative number of simulated hits at *X* = 64*, N* = 1, 000 for all values of *p* in the interval [0, 0.04]. The interval of values of *p* in [*L*_0.025_*,U*_0.975_] = [0.0032, 0.0377] is responsible for 95% of the simulated hits at *X*.

**Figure 3:**
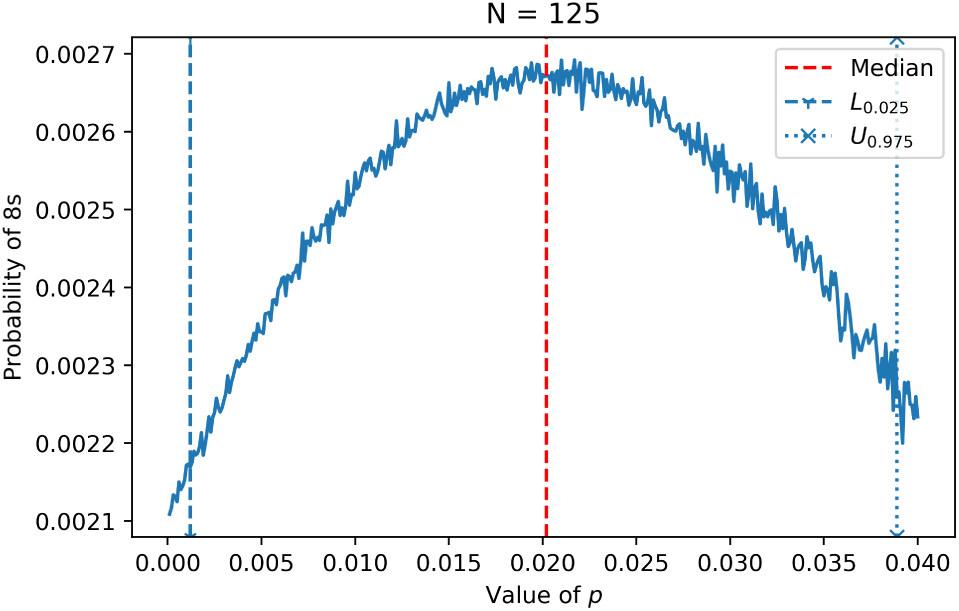
A histogram of the relative number of simulated hits at *X* = 8*, N* = 125 for all values of *p* in the interval [0, 0.04]. The interval of values of *p* in [*L*_0.025_*,U*_0.975_] = [0.0012, 0, 0389] is responsible for 95% of the simulated hits at *X*.

**Discussion of findings** The three histograms depicted in Figures 1, 2 and 3 share a few common qualitative features. First, they appear single peaked and rather symmetric. Recall furthermore that

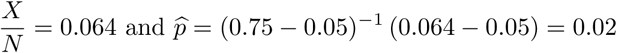

Observe that the median and the modus change only very slightly over the three histograms, We obtain the following ranking (for each case studied in this subsection)

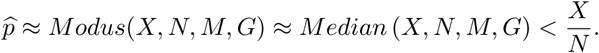

Furthermore, we find

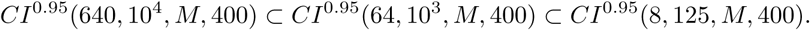

The effect on the size of the confidence intervals is significant. The size of the corresponding interval for *N* = 1, 000 is more than double the size for that for *N* = 10, 000, whereas the confidence interval for *N* = 125 is almost three times the latter size.

### 2.4 Results for 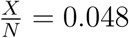*, G* = 400 and *N* ∈ {10^4^, 10^3^, 125}

**Figure 4:**
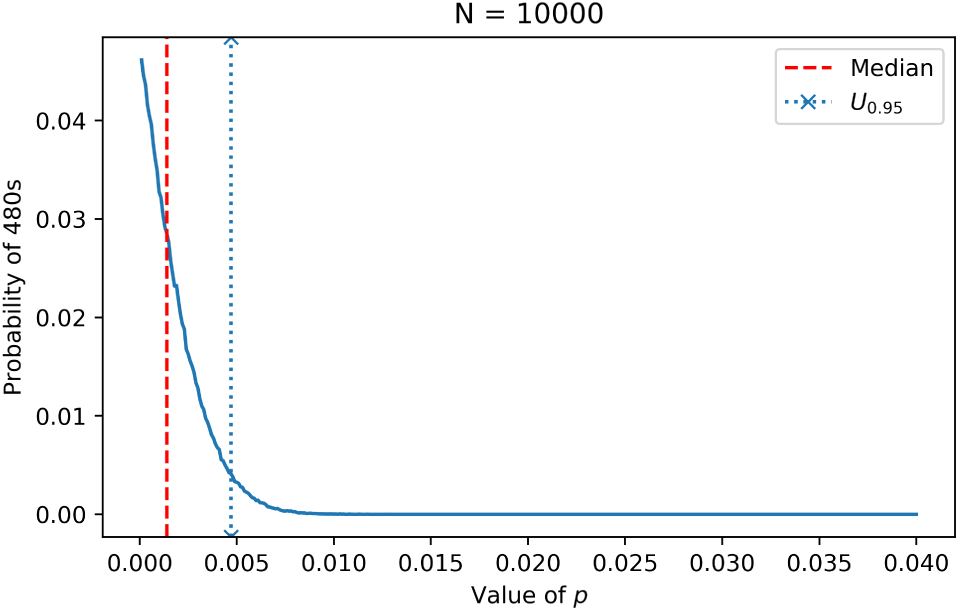
A histogram of the relative number of simulated hits at *X* = 480*, N* = 10, 000 for all values of *p* in the interval [0, 0.04]. The interval of values of *p* in [0*, U*_0.95_] = [0, 0.0047] is responsible for 95% of the simulated hits at *X*.

**Figure 5:**
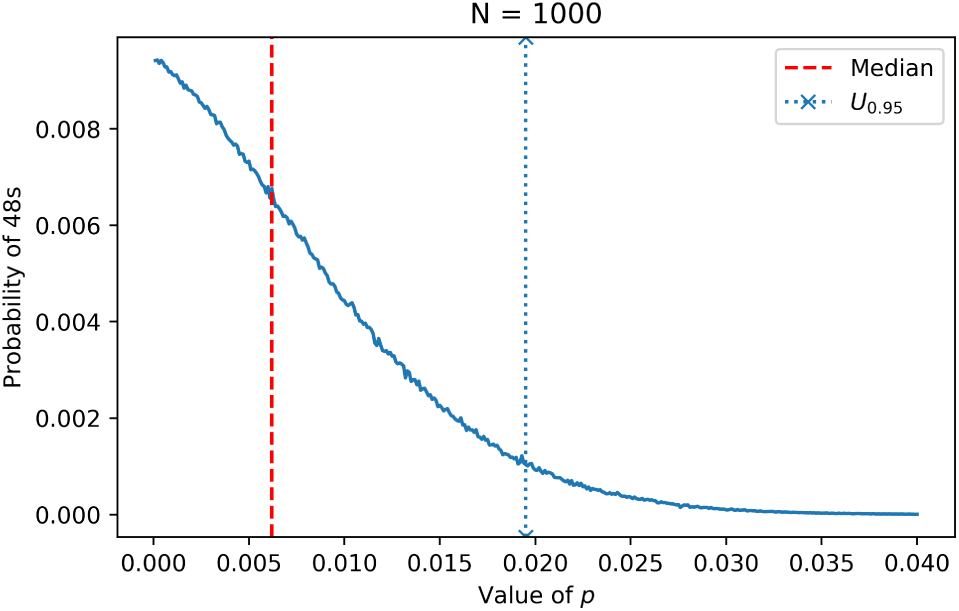
A histogram of the relative number of simulated hits at *X* = 48*, N* = 1, 000 for all values of *p* in the interval [0, 0.04]. The interval of values of *p* in [0*, U*_0.95_] = [0, 0195] is responsible for 95% of the simulated hits at *X*.

**Figure 6:**
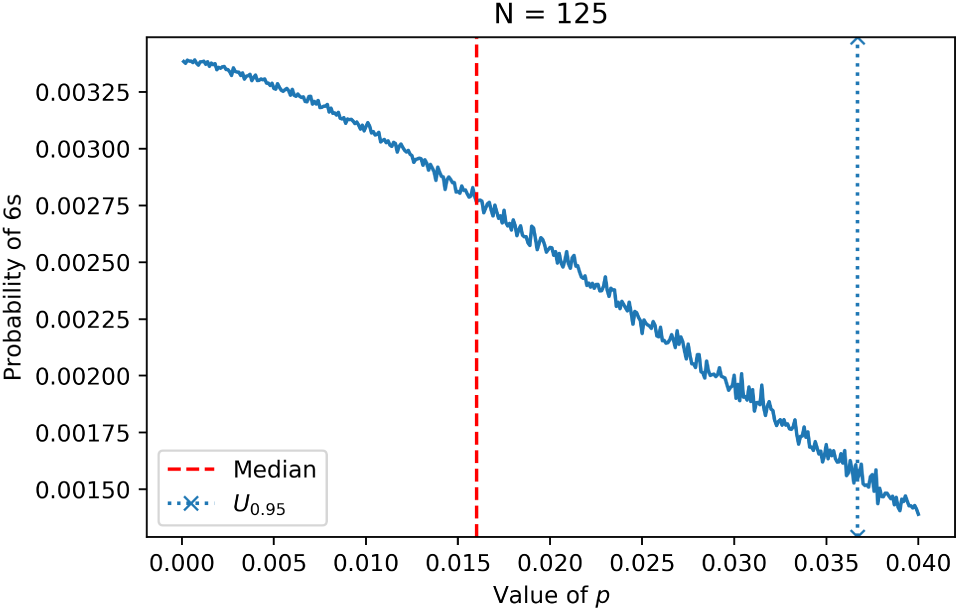
A histogram of the relative number of simulated hits at *X* = 6*, N* = 125 for all values of *p* in the interval [0, 0.04]. The interval of values of *p* in [0*, U*_0.95_] = [0, 0.0367] is responsible for 95% of the simulated hits at *X*.

**Discussion of findings** Figures 4, 5, and 6 share a few common qualitative features, but differ strikingly from the three histograms of the previous subsection. First, these histograms are far from symmetric, they appear single peaked at zero. Furthermore,

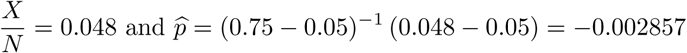

Observe that the modus changes only very slightly over the three histograms, if at all, but equals zero. The median for the three cases is positive, it shifts considerably and the higher *N* is the closer the median gets to zero. This seems quite intuitive, as unlikely results in the sense that 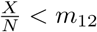, should occur less and less frequently if the sample size increases. We obtain the following ranking (for each case studied in this subsection)

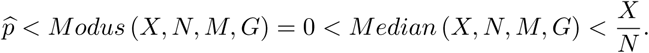

The modus appears to be at zero, which will simply not do as a point estimator of the prior. It is logically inconsistent to have positives if the prior is truly equal to zero.

For the confidence interval we find

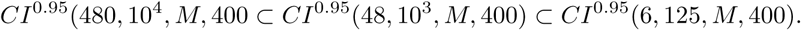

Again, in line with intuition, we see that for larger *N*, keeping the ratio 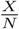 fixed, the size of the confidence interval shrinks.

### 2.5 Results for 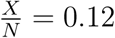*, G* = 400 and *N* ∈ {10^4^, 10^3^, 125}

**Figure 7:**
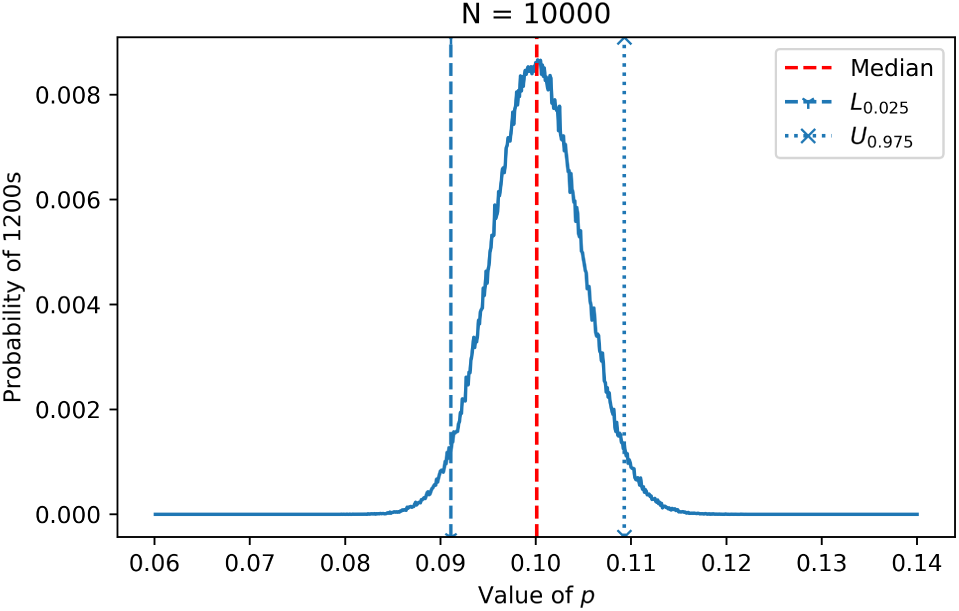
A histogram of the relative number of simulated hits at *X* = 1200*, N* = 10, 000 for all values of *p* in the interval [0, 0.02]. The interval of values of *p* in [*L*_0.025_*,U*_0.975_] = [0.0911, 0.1093] is responsible for 95% of the simulated hits at *X*.

**Figure 8:**
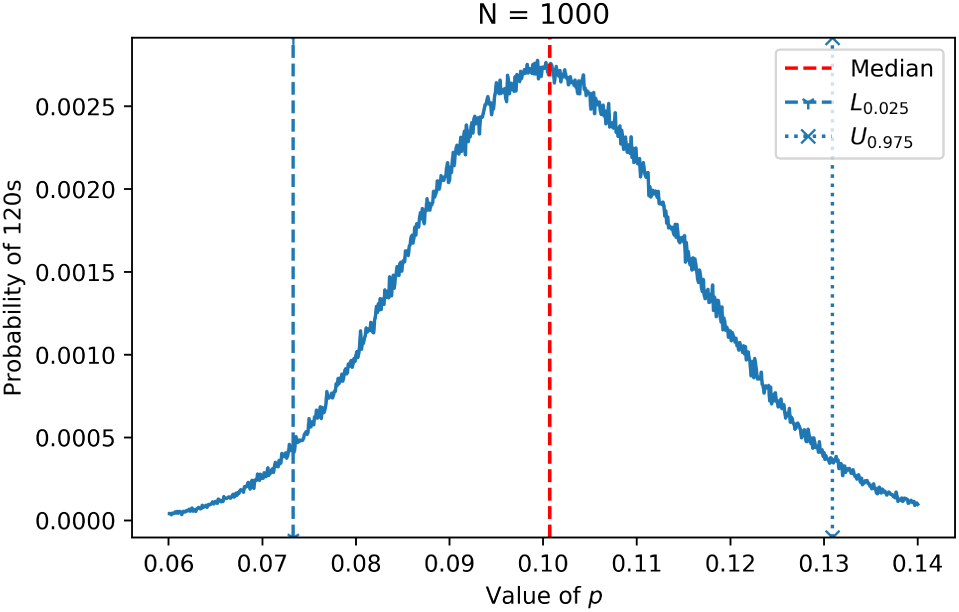
A histogram of the relative number of simulated hits at *X* = 120*, N* = 1, 000 for all values of *p* in the interval [0, 0.02]. The interval of values of *p* in [*L*_0.025_*, U*_0.975_] = [0.0733, 0.1309] is responsible for 95% of the simulated hits at *X*.

**Figure 9:**
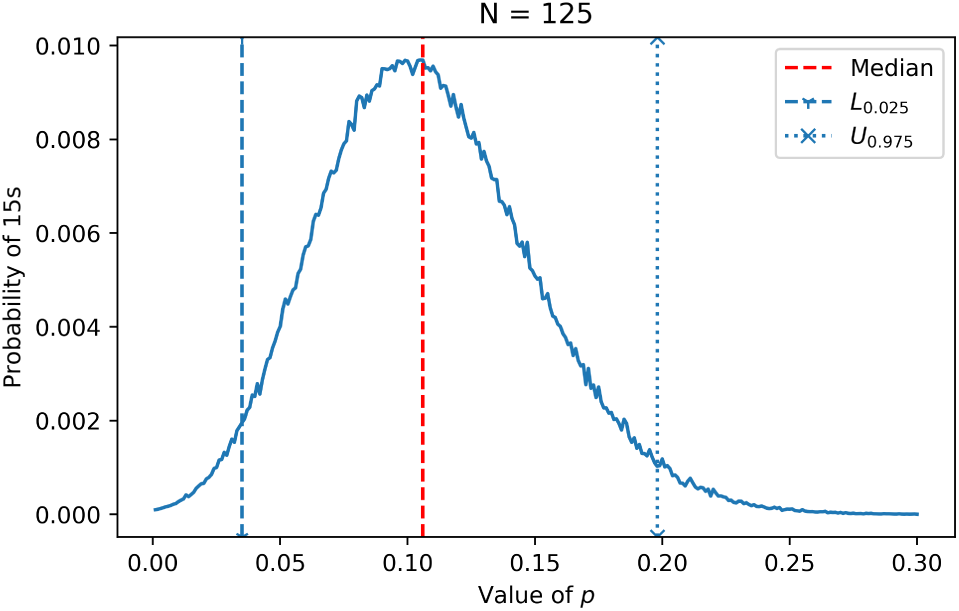
A histogram of the relative number of simulated hits at *X* = 15*, N* = 125 for all values of *p* in the interval [0, 0.3]. The interval of values of *p* in [*L*_0.025_*,U*_0.975_] = [0.0351, 0.1984] is responsible for 95% of the simulated hits at *X*.

**Discussion of findings** The figures in this subsection share a few common qualitative features, but the first two seemingly share more qualitative features among them and with the first set of three histograms, than with the third histogram. Again the histograms appear single peaked, the first two seem rather symmetric, the last one seems skewed.

The median and the modus appear quite close in the first two figures. Furthermore, we have

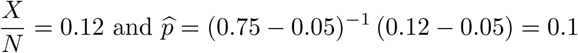

Observe that the median and the modus change only very slightly among the three histograms. We obtain the following ranking (for each case studied in this subsection)

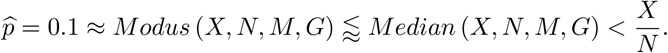

Furthermore, we find

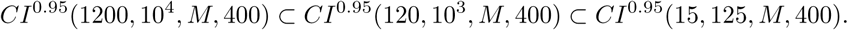

Observe that the median again changes only very slightly over the three histograms, but the confidence intervals change tremendously in size.

## 3 Conclusion

For the first couple of weeks as the Corona crisis developed, we have been merely bewildered spectators at the side line, wondering how to make sense of phenomena with relevant data and estimates lacking universally. Frankly, we questioned the validity of many of the statements made by scientists, politicians and serious media. Quite recently we found an opportunity to make constructive use of our experience in designing Monte Carlo simulations for problems in which analytical distributions of relevant phenomena are very hard to obtain. We designed a tool^9^ to find base rates underlying certain tests.

Actually, we set out on a larger idea of which this is the first preliminary paper^10^. We propose a procedure based on Monte Carlo simulation based analysis with inputs: a sample of *N* from a certain population is taken, *X* is the number of positives and *M* is the matrix combining the reliability of the test, i.e.,

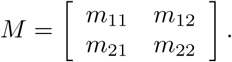

This matrix satisfies 1 = *m*_11_ + *m*_21_ = *m*_12_ + *m*_22_, where *m*_11_ may be called the true positive rate, *m*_21_ is the false negative rate (or type II error rate), *m*_12_ is the false positive rate (or type II error rate) and *m*_22_ is the true negative rate.

We may distinguish several point estimators for the base rate *p* of certain populations, and the following two are seemingly^11^ frequently used:

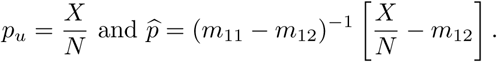

The subscript *u* stands for ‘unadjusted.’ The first estimator has been used in recent studies (e.g., Bendavid *et al*. [2020]) as a quick-fire solution disregarding test reliabilities, the second should however be considered as a slightly more precise point estimator incorporating the probabilities of false positives in the test. We have the following rankings among those two estimators:

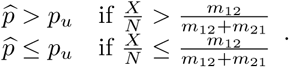

So, only by sheer ‘luck’ both estimators coincide in general. Furthermore,

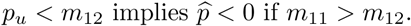

In this paper we add three new estimators of the base rate in a population. Two are point estimators, the third is an interval estimator, or confidence interval. We must stress that for the present procedure we assume the matrix *M* to be deterministic.

The **modus** is the smallest prior which yielded the highest number and hence proportion of positives equal to *X* in our MC simulation for sample size *N* using *deterministic* reliability matrix *M*. The **median** is the upper bound set of ranked priors starting at the lowest value, responsible for (approximately) half of the simulated hits equal to *X* in the MC simulation. We interpret a our (1 − *α*)-confidence interval among the priors as that it gives us the set of priors accounting for a proportion of 1 − *α* of outcomes yielding *X* hits in the Monte Carlo simulation.

We focus on the following findings regarding this collection of point and interval estimators. By elimination of alternatives, the final bullet point gives the most preferred pair of estimators, in our opinion.

- In many cases the median, modus and 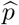 are quite close, and are to be found rather central in any standard two-sided confidence interval.
- Confidence intervals shrink in size as the number *N* increases, i.e., the discriminatory power of the procedure increases in the usual manner.
- The median is always in the range of the most used confidence intervals (90%, 95%, 99% two-sided).
- The sample size *N* has negligeable influence on the median, the modus and 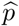 relative to the size of the corresponding two-sided 95%-confidence intervals generated, provided that the resulting histogram is **close to symmetric**. So, **rather small samples may provide rather reliable estimators for cases yielding symmetric histograms**.
- It may happen that 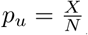 does not fall into the two-sided 95%-confidence interval of the procedure (cf., e.g., Figures 1 − 7). This rules out this estimator as a universally applicable point estimator, in our opinion.
- It may happen that 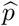 is negative, which rules this estimator out as a universally applicable point estimator by logic.
- It may happen that the modus is equal to zero (cf., e.g., Figures 4 − 6), which rules out the modus as a universally applicable point estimator by logic.
- The sample size *N* is of significant influence on the median and of no influence on the modus and 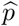 (as the latter are smaller than or equal to 0) for low ratios of 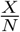. The median decreases considerably if *N* is increased.
- Both the median and the confidence intervals universally make sense as concepts, as well as as estimators.

## 4 Appendix: the procedure applied to two data points from a recent study

On Saturday April 18, while trying to finalize this preliminary paper, we found a study reporting on tests in the county of Santa Clara in California (Bendavid *et al*. [2020]). We gladly refer to the paper for more details of this interesting (also) preliminary report.

In a rather precisely described case, the authors found a number of 50 positives in a test of size 3330. So, for the first two inputs necessary necessary, we took *X* = 50 and *N* = 3330. Determining *M*, the matrix summarizing the test reliability was a little bit more problematic for us. The authors provided a lot of numbers regarding the test validity which are highly relevant to our framework, but frankly, we were a quite dazzled by them. We took the liberty of generating the following matrix of test reliability (the underlying numbers were found in Bendavid *et al*. [2020]) under the presumption that this is indeed what the authors intended for the unadjusted case:

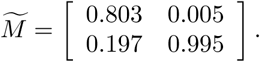

This matrix was obtained by interpreting the statement:

… provides us with a combined sensitivity of 80.3% (95 CI 72.1–87.0%) and a specificity of 99.5% (95 CI 98.3–99.9).

Following standard practice, we took 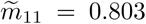 and 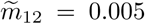 which immediately induces all four entries in the reliability matrix.

### 4.1 Findings

We ran our procedure^12^ using these numbers and obtained results visualized in Figure 10. We interpret the least sophisticated framework, i.e., we do the rough estimation on total population level, which happens to yield the lowest valued estimator of all estimators of the base rate presented in Bendavid *et al*. [2020].

**Figure 10:**
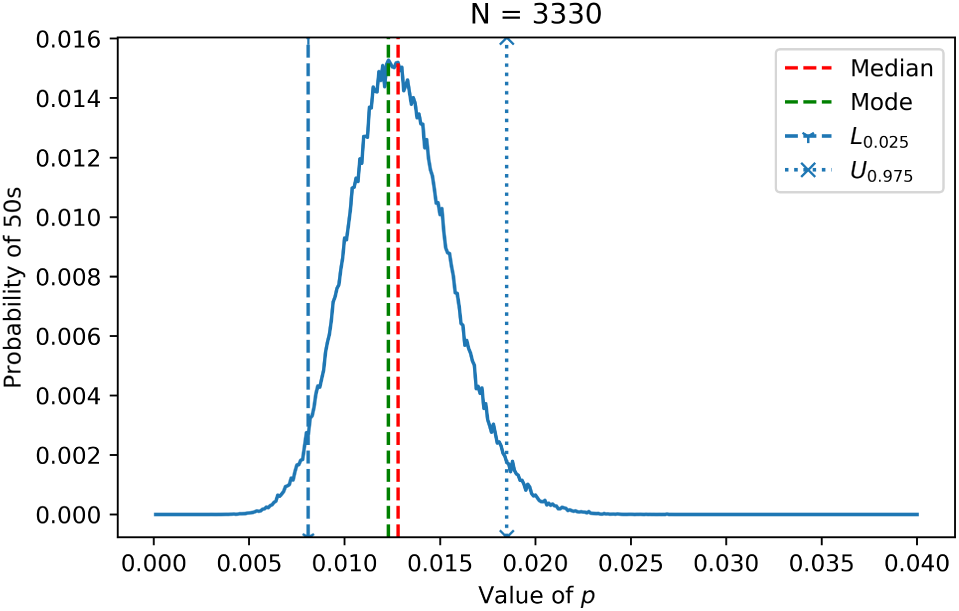
The output of our Monte Carlo simulation based procedure obtained from our interpretation of the reliability matrix in Bendavid *et al*. [2020] applied to the aggregate findings. The median, the modus and the 95%-confidence interval are indicated.

Figure 10 is rather illustrative on its own, but for the reader’s convenience we summarize some relevant candidate estimators below.

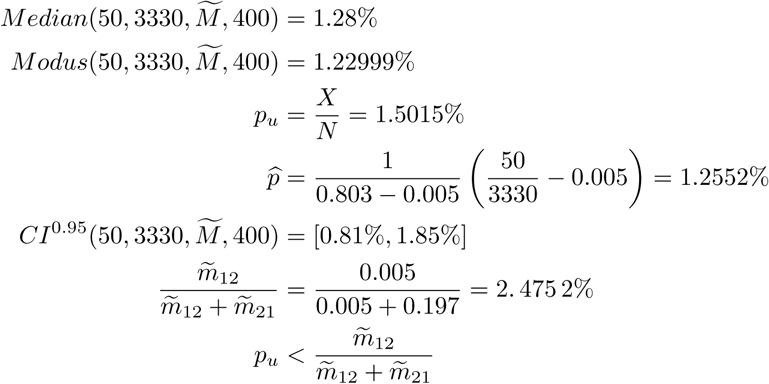

Clearly, we have

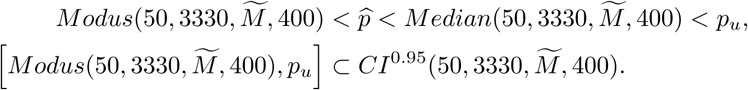

Hence, all point estimators are in the 95%-confidence interval. The upper bound of the confidence interval tells us that the priors exceeding this upper bound account for less than 0.025% (combined) of the hits equal to 50 in the Monte Carlo simulation.

The modus and median are rather close and located rather centrally in the 95%-confidence interval. Based on our preferences we would recommend using *Median*(50, 3330*, M, G*) = 1.28% as the point estimator and *CI*^0.95^(50, 3330*, M, G*) = [0.81%, 1.85%] as a reasonable confidence interval.

### 4.2 Comments

On the one hand, the unadjusted point estimator is found to be

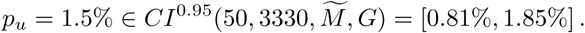

So, since this estimator is in our 95%-confidence interval, we do not reject the point estimator *p_u_*.

On the other, this number might be a bit on the high side as we have shown in the concluding section of this paper. Even without having computed 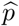 we know

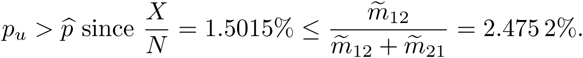

Indeed, the realization of the latter estimator was 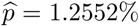. If we compare the latter estimator with the concepts introduced in the body of this paper we see

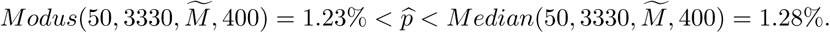

This might indicate that we should expect the true value to be closer to the threesome mentioned than to *p_u_*.

Our confidence interval is obtained directly from the Monte Carlo simulation with inputs 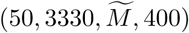. Bendavid *et al*. [2020] report the following two confidence intervals

#### [1.11%, 1.97%] and [1.07%, 1.93%]

Under the reservation that we might not be comparing the same objects, our confidence interval is larger than any of the pair mentioned, located more to the left, moreover the upper and lower bounds are lower than the corresponding bounds of the pair they mention.

Note finally, none of the three alternative point estimators can be rejected for either confidence interval presented by Bendavid *et al*. [2020] pertaining to their unadjusted prior either, as clearly the estimators are in the intersection of both, i.e.,

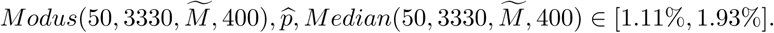

## Data Availability

https://github.com/abhishta91/antibody_montecarlo

## Footnotes

1 In this paragraph we are not overusing the word approximately. Each use of the word is intentional.

2 https://it.businessinsider.com/esclusiva-cosa-rivelano-i-primi-test-di-robbio-primo-paeseitaliano-a-fare-i-test-sullimmunita-a-tutti-i-cittadini/

3 At the moment of writing a problem in the Netherlands. The Dutch government had the aim of testing 17,000 people per day from a certain date onwards, but this date has gone by and the maximum daily number of tests taken in reality is approximately 7,000.

4 See https://github.com/abhishta91/antibody_montecarlo

5 The number of True Positives.

6 The number of True Positives tested positive.

7 The number of True Negatives tested negative.

8 The number of True Negatives tested positive, i.e., the so-called false positives.

9 Due to time pressure, we did a hasty check on literature. So, none of this line of thinking/modeling might be new, and we apologize for wasting your time. However, our sincere intention was to offer some help.

10 The second paper, to appear in a couple of days, proceeds on this one, but will take another hurdle in estimating base rates, namely the real-life problem of test reliability matrices which are estimates themselves (hence, with all components being stochastic).

11 Seemingly, because none of the reports we found use explicit formulas. Recalculating one of the reported numbers in Bendavid et al. [2020] yields a perfect match. In a report (in German) by Streeck et al. [2020] only specificity *m*_22_ *>* 0.99 is mentioned which bounds *m*_12_, but not *m*_11_. Taking both specifity and sensitivity equal to 99% yields an outcome which is compatible with their estimation.

12 The Python code may be found at https://github.com/abhishta91/antibody_montecarlo

